# Image-based computational hemodynamics analysis of systolic obstruction in hypertrophic cardiomyopathy

**DOI:** 10.1101/2021.06.02.21258207

**Authors:** Ivan Fumagalli, Piermario Vitullo, Roberto Scrofani, Christian Vergara

## Abstract

Hypertrophic Cardiomyopathy (HCM) is a pathological condition characterized by an abnormal thickening of the myocardium. When it affects the medio-basal portion of the septum, it is named Hypertrophic Obstructive Cardiomyopathy because it induces a flow obstruction in the left ventricle outflow tract, which may compromise the cardiac function and possibly lead to cardiac death. In this work, we investigate the hemodynamics of different HCM patients by means of computational hemodynamics, aiming at quantifying the effects of this pathology on blood flow and pressure gradients and thus providing clinical indications that may help diagnosis and the design of surgical treatment (septal myectomy). To this aim, we employ an enhanced version of an image-based computational pipeline proposed in a previous work, integrating fluid dynamics simulations with geometrical and functional data reconstructed from standard cine-MRI acquisitions. Blood flow is modelled as an incompressible Newtonian fluid, The corresponding Navier-Stokes equations are solved in a moving domain obtained from cine-MRI, whereas the valve leaflets are accounted for by a resistive method.

## INTRODUCTION

Hypertrophic Cardiomyopathy (HCM) is a typically congenital cardiac disorder characterized by an abnormal thickening of the myocardium (wall thickness exceeding 15 mm) without additional causes that may induce secondary hypertrophy [1]. It has a prevalence of 0.2–0.6% in the western world and an overall annual mortality rate of 1%. When the thickening affects the medio-basal region of the interventricular septum, this condition may cause a flow obstruction in the Left Ventricle Outflow Tract (LVOT), and thus takes the name of Hypertrophic Obstructive Cardiomyopathy (HOCM). People with HOCM often remain oligosymptomatic or even asymptomatic for many years. Anyway, they may develop dyspnoea, angina pectoris, or stress-induced syncope, with an increased risk of sudden cardiac death, particularly in young people and athletes, and its pathological effects can be worsened by concurrent conditions increasing the ventricle afterload, such as hypertension or aortic stenosis. The LVOT obstruction is dynamic and largely influenced by changes in left ventricular loading and contractility, with subsequently increased systolic pressure in the Left Ventricle (LV) and possible secondary mitral regurgitation, myocardial ischemia, and reduction in cardiac output. In particularly severe cases, a Systolic Anterior Motion (SAM) of the mitral valve can be induced, thus further worsening the LVOT obstruction [2], [3], [4], [5].

One of the most widely employed surgical treatment for a pathologically relevant HOCM is septal myectomy, namely the resection of a portion of the septum, in order to recover a physiological patency of the LVOT [1]. Therefore, identifying the location and extension of the septal region inducing the obstruction is of paramount importance in the clinical decision on the surgical treatment and the preoperative design of the procedure.

In order to obtain quantitative indications on blood flow, pressure gradients and wall shear stresses associated to it, computational hemodynamics approaches have proved to be extremely helpful, thanks to their flexibility and level of detail. In this regard, two main standpoints are currently adopted: Fluid-Structure Interaction (FSI) simulations and prescribed-motion Computational Fluid Dynamics (CFD). The first approach, employed for example in [6], [7], [8], [9], [10], [11], consists in the coupled solution of the fluid dynamics of blood flow and of the structure mechanics of the myocardium and valves, thus entailing a possibly very high computational cost. On the other hand, in image-based CFD, the displacement of the myocardium and valves leaflets is reconstructed from kinetic medical images and then prescribed as endocardial data to obtain the fluid domain configuration. This latter approach, that reduces the mathematical complexity of the problem with respect to a full FSI system, at the expense of a more complex image processing procedure, has been shown to provide insightful indications on cardiovascular pathologies in a series of works, including the computational study of HCM and LV hemodynamics as in [12], [13], [14], [15].

The goal of the present work is to investigate the hemodynamics in the systolic phase and the LVOT obstruction severity and extent in patients suffering from HCM, and to provide quantitative indications that may help to assess the severity of the condition and to design the possible surgical treatment. To this aim, we start from the computational pipeline proposed in [15], based on cine-MRI data, and we introduce further improvements based on the integration among short-axis and long-axis views, to accurately reconstruct the patient-specific geometry and motion of the LV and of the ascending aorta and to set up the conditions for a CFD analysis. This motion is then extended to the mitral valve leaflets, which are immersed in the fluid domain by a resistive method [15], [16].

## METHODS

In the present section, we describe the imaging data on which this work is based, and we introduce a computational pipeline encompassing image processing, surface morphing, and numerical simulations, for the study of the hemodynamics in the left ventricle (LV) and the ascending aorta.

### Patient data

Cine-MRI data of three patients were provided by L. Sacco Hospital in Milano, with the approval of the Ethics Committee and accordingly to the ethics guideline of the institutions involved, including the signed consent of the patient. For each patient, the data comprise different series, with different resolution properties: a volumetric short-axis acquisition, with a spacing and a slice thickness of 8mm along the LV main axis, a space resolution of 1mm and a time resolution of 1/20s; a set of single-slice, two-dimensional long-axis acquisitions on the so-called *two-chambers, three-chambers* and *four-chambers* planes, with space resolution of 1mm and time resolution that may be of 1/20s or 1/30s. We point out that these are standard cine-MRI data, that are routinely acquired during diagnostical procedures: the computational pipeline presented in this work does not require the setup of *ad-hoc* acquisitions. Two of these patients (named in the following as Patient 1 and Patient 2) have never been studied from a computational standpoint, whereas the last one (Patient 3 from now on) was the subject of an investigation on systolic anterior motion [15], and it is reported in this broader study both for comparison and in order to analyse the HOCM-induced obstruction separately from the effects of SAM.

### Reconstruction of geometry and motion

We devised a computational procedure, encompassing the processing of images and surfaces and the generation of surface and volumetric computational meshes. Based on standard cine MRI, we developed an algorithm to merge all the information coming from the different acquisition series. Indeed, as described above, the only three-dimensional data is represented by the short-axis series, which however has a relatively coarse resolution along the LV main axis. By proper interpolation, we could enhance the short-axis images with the long-axis acquisitions, obtaining an *artificial*, time-dependent series of volumetric images with a uniform space resolution of 1mm in all directions. The algorithm was implemented in Matlab, and further details on it can be found in [17].

Thanks to these artificial images, we could improve the reconstruction procedure presented in [15], resulting in the following reconstruction steps:

1. for each instant of the artificial image, we segment both the endocardium and the epicardium of the LV;
2. intersecting the endocardium and epicardium surfaces obtained at the previous step, we obtain a volumetric representation of the myocardium as a level-set image;
3. we choose the end of systole as a reference configuration, and we apply a registration algorithm among the level-set images of the myocardium, in order to obtain a displacement field for each acquisition time;
4. we apply the displacement field to the endocardium surface of the reference configuration to obtain its evolution throughout the heartbeat;
5. a template geometry of the aorta provided by Zygote [18] is adapted to the patient-specific LV, based on its aorto-mitral annulus, in order to obtain a complete computational domain for the study of the systolic phase;
6. the computational volumetric mesh is then automatically generated, and the displacement fields are stored for the purposes of moving-domain CFD simulations.

We point out that the main difference with respect to the procedure employed in [15] is the generation of the enriched artificial image: on its basis we could accurately capture the shortening and elongation of the LV directly from segmentation, without the need to measure it separately, and also the patient-specific aorto-mitral annulus that is exploited at point 5. As reconstruction tools, we employed the Medical Image Toolkit (MITK) [www.mitk.org], [19], [20] for the segmentation step, a procedure based on SimpleITK [simpleitk.org] for registration, and for the other steps we employed the tools presented in [21] and other ad hoc semi-automatic tools hinging upon the Visualization Toolkit (VTK) [vtk.org] and the Vascular Modeling Toolkit (VMTK) [vmtk.org], [22].

In order to complete the geometrical description of the domain, we adapted a template mitral valve geometry provided by Zygote to the patient-specific reconstructed geometry: usually, standard MRI acquisitions are not sufficiently detailed to allow an accurate reconstruction of valves. Since we are going to investigate only the systolic phase, and no mitral regurgitation is reported for any of the patients, the valve is kept closed during the whole systolic phase, while moving along with the ventricle. We point out that, in order to study the effects of the sole hypertrophy on the flow, we consider a healthy valve also for Patient 3, neglecting SAM.

### Mathematical model – Computational Fluid Dynamics

Under the common assumption that blood is an incompressible Newtonian fluid, at least in the heart chambers and in large vessels [23], we can model its flow by incompressible Navier-Stokes equations, with density ρ = 1.06 · 10^3^ kg/m^3^ and viscosity µ = 3.5 · 10^−3^ Pa · s. An Arbitrary Lagrangian-Eulerian (ALE) formulation of the equations allows to prescribe the domain displacement reconstructed in the previous section, whereas the effects of the mitral valve on the flow is accounted for by the Resistive Immersed Implicit Surface (RIIS) method. This method, laying in the general class of immersed boundary and fictitious domain techniques, was introduced in [16], based on the Resistive Immersed Surface method developed in [24], [25], and it has been applied to the study of the Systolic Anterior Motion (SAM) of the mitral valve in [15]. We focus our study on the systolic evolution, and since none of the patients under investigation presented anomalies in their systemic pressure, as a boundary condition we consider a physiological evolution of the aortic pressure, derived from Wiggers diagrams [26]. The problem is numerically solved by using a first order semi-implicit time discretization and the Finite Element Method for space discretization, by means of life^x^ [https://gitlab.com/lifex/lifex], a multiphysics high-performance library based on the deal.II core [https://www.dealii.org] and developed in the iHEART project ^1^. For all the patients, we discretize the domain by a hexahedral computational mesh with an average size of h = 1mm and are a local refinement to h = 0.3mm in the region of the mitral valve and the LVOT. We performed a merge convergence test to ensure that no significant differences may be found by using a finer mesh. A simulation timestep of Δt=10^−4^ s was adopted, and a smooth spline interpolation was used to represent the reconstructed displacements on the simulations time grid.

### Outputs of interest

The reconstruction pipeline and the computational methods described above allow to obtain different relevant outputs about the overall cardiac function and the hemodynamics of the patients under investigation. In addition to well-known biomarkers such as the stroke volume (SV), the end-diastolic/end-systolic volume (ESV/EDV) and ejection fraction (EV), an accurate reconstruction of the LV volume evolution throughout the heartbeat is recovered. Then, focusing on the systolic phase, we can assess the possible HCM-induced obstruction in terms of velocity distribution, aortic jet development and pressure gradients. A particularly relevant indicator for the clinical assessment of the obstruction severity is the distribution of pressure along the septum. This information can help the surgeon in the design of the most common treatment for HOCM, namely septal myectomy, because it provides a quantitative measure of the spatial location and extension of the obstruction.

## RESULTS

In this section, we present the outcomes of the application of the proposed computational pipeline to the data of three patients described above.

From the reconstruction procedure we obtain the displacement fields shown in Figure 1. We can notice how the distribution of displacement and its intensity are significantly different among the three patients under investigation, expressing the high variability of the effects of HCM on the ventricle contraction. For example, if for all three we observe a generally reduced motion of the septal wall, we can also notice that for Patient 1 this reduction is mostly concentrated in the region near to the ventricular base, whereas Patient 2 has a more homogenously distributed hypokinesis. These differences in the movement of the endocardial wall are reflected in the evolution of the volume of the LV cavity, presented in Figure 2. A remarkable difference in this sense is displayed by Patient 2, for which we can appreciate a slowed down diastolic expansion after the systolic peak at 0.4s. This is in accordance with the reduced ejection fraction of Patient 2, as we reported in the table of Table 1. From the same table, we can compare the volume measurements obtained by our reconstruction with those estimated during the data acquisition. Such clinical estimates are based on an approximation of the LV as an ellipsoid, which may not be particularly accurate for the end systolic volume (ESV), especially in the case of hypertrophic patients for which contraction may be spatially inhomogeneous. On the other hand, we notice that end diastolic volume (EDV) values show good agreement between the reconstructed and estimated values, with a relative difference of less than 8% for all patients.

**Table 1.** End-diastolic volume (EDV), end-sytolic volume (ESV) and ejection fraction (EF) as reconstructed from the computational procedure (above) and from clinical estimations (below).

| PT | EDV [ML] | ESV [ML] | EF [%] |
| --- | --- | --- | --- |
| 1 | 138 | 68 | 51 |
| 2 | 162 | 94 | 42 |
| 3 | 102 | 43 | 57 |
| 1 | 128 | 81 | 37 |
| 2 | 152 | 103 | 32 |
| 3 | 101 | 37 | 63 |

**Figure 1.**
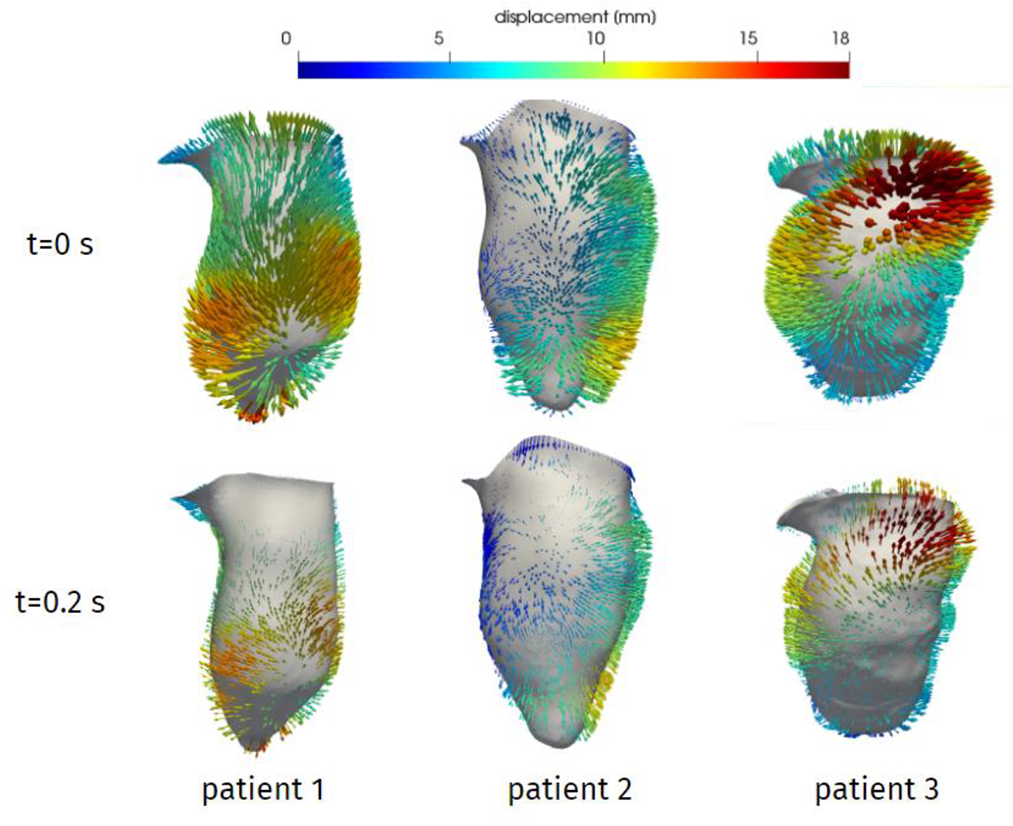
Reconstructed displacement field of the three patients, at the end of diastole (t=0 s) and in late systole (t=0.2s). LV aligned vertically, with septal wall on the left.

**Figure 2.**
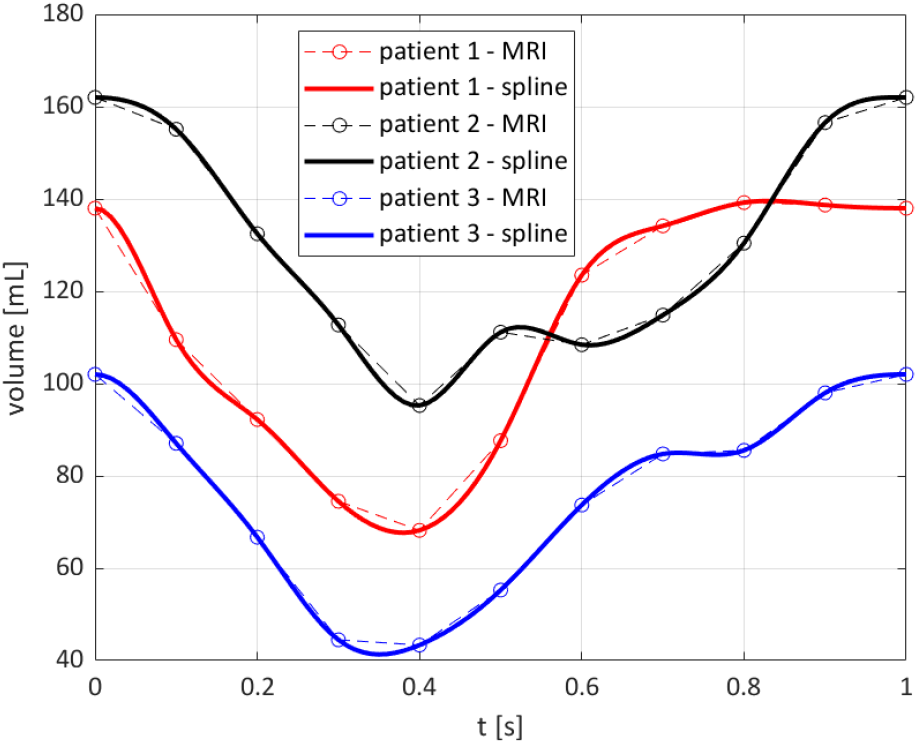
Evolution of the reconstructed volume of the LV cavity.

After assessing the reconstruction procedure and its outcomes in terms of geometry and displacements, we now present the results of the computational hemodynamics simulations under such prescribed motion. We remind that, since the focus of the present work is to investigate the possible obstructions induced by HCM on the flow, we focus on the systolic phase. In Figure 3 we can see the blood velocity field during systole for the three patients under investigation. Regarding the velocity peak, we can see that it is reached at different times for the three patients. Patient 1 shows a very quick blood acceleration in the early stages of the ejection, with a strong jet involving the whole aortic root and a maximum velocity of 1.38 m/s attained at t=0.08 s, then followed by a relatively slower deceleration. Patient 2 and 3, instead, present a more progressive development of the aortic jet, with a maximum velocity of 1.31 m/s and 3.0 m/s attained at t=0.23s and 0.20 s, and high velocity values (>1 m/s) can be appreciated in the aortic root also at later stages of the systolic phase. In terms of velocity peak, Patients 1 and 2 lay just above the range of physiological values (1-1.2 m/s), whereas Patient 3 is definitely in the pathological range.

**Figure 3.**
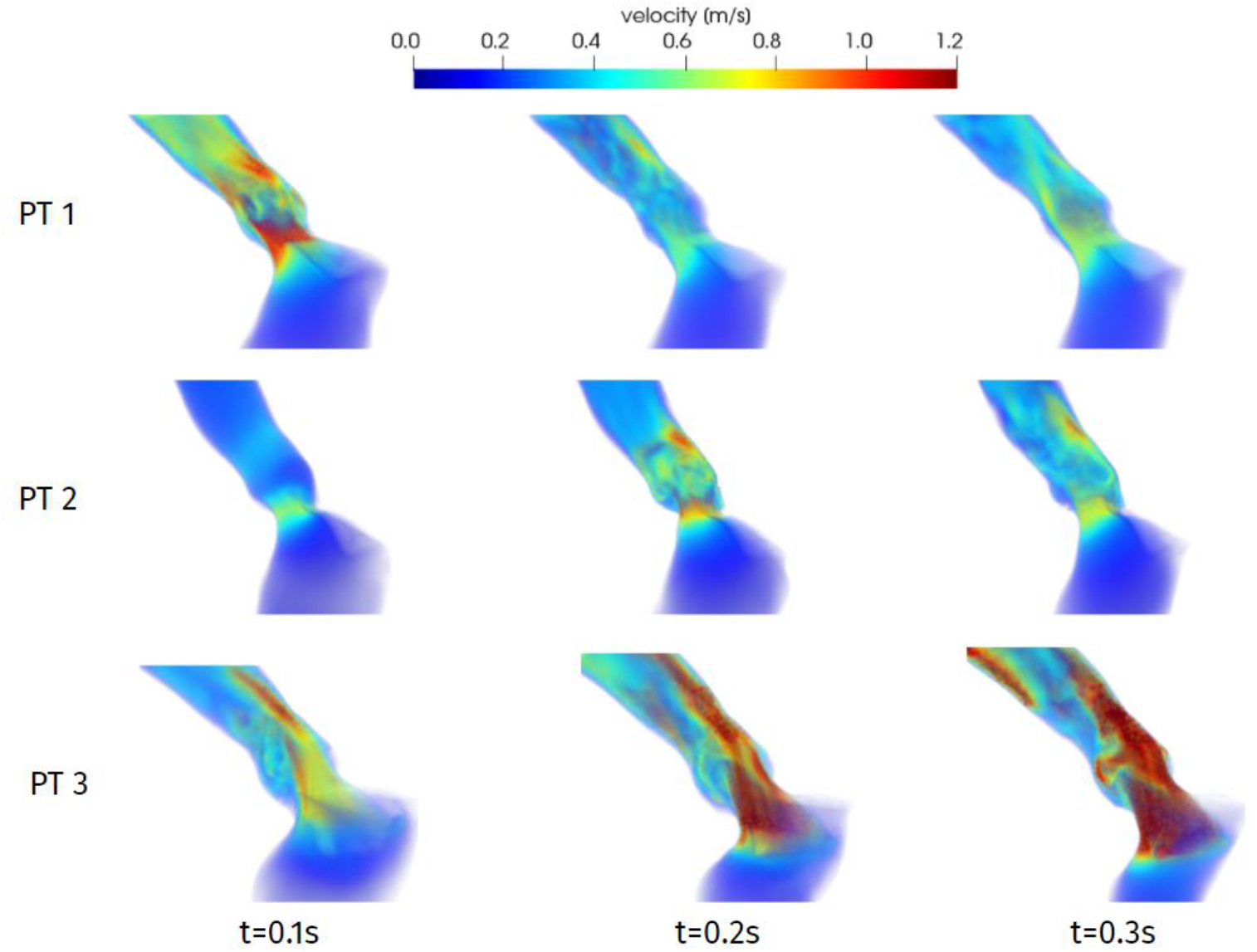
Velocity distributions at significant times during systole.

One of the main quantities that is inspected in the assessment of the hypertrophy-induced obstruction is the intraventricular pressure gradient, that is the variation of pressure in the LVOT. In order to evaluate it, we compute the difference Δp=p-p_AO_ between the pressure p and its value p_AO_ in the centre of Valsalva sinuses, and in Figure 4 we display the distribution on a two-dimensional longitudinal slice. For all patients and at all times, we notice that significant pressure gradients can be appreciated only in the LVOT and (at a smaller extent) in the Valsalva sinuses, whereas pressure is essentially uniform elsewhere. Regarding the differences among the patients, the overall pressure gradient of Patient 1 is always between -3 and 3 mmHg, whereas for the other two the gradient reaches more than double such values. Regarding time evolution, we can notice that each patient experiences an increase or a decrease of Δp in different stages of the systole, reflecting the variability that we already observed in the comments above about the time dependence of the displacements and volume of the LV.

**Figure 4.**
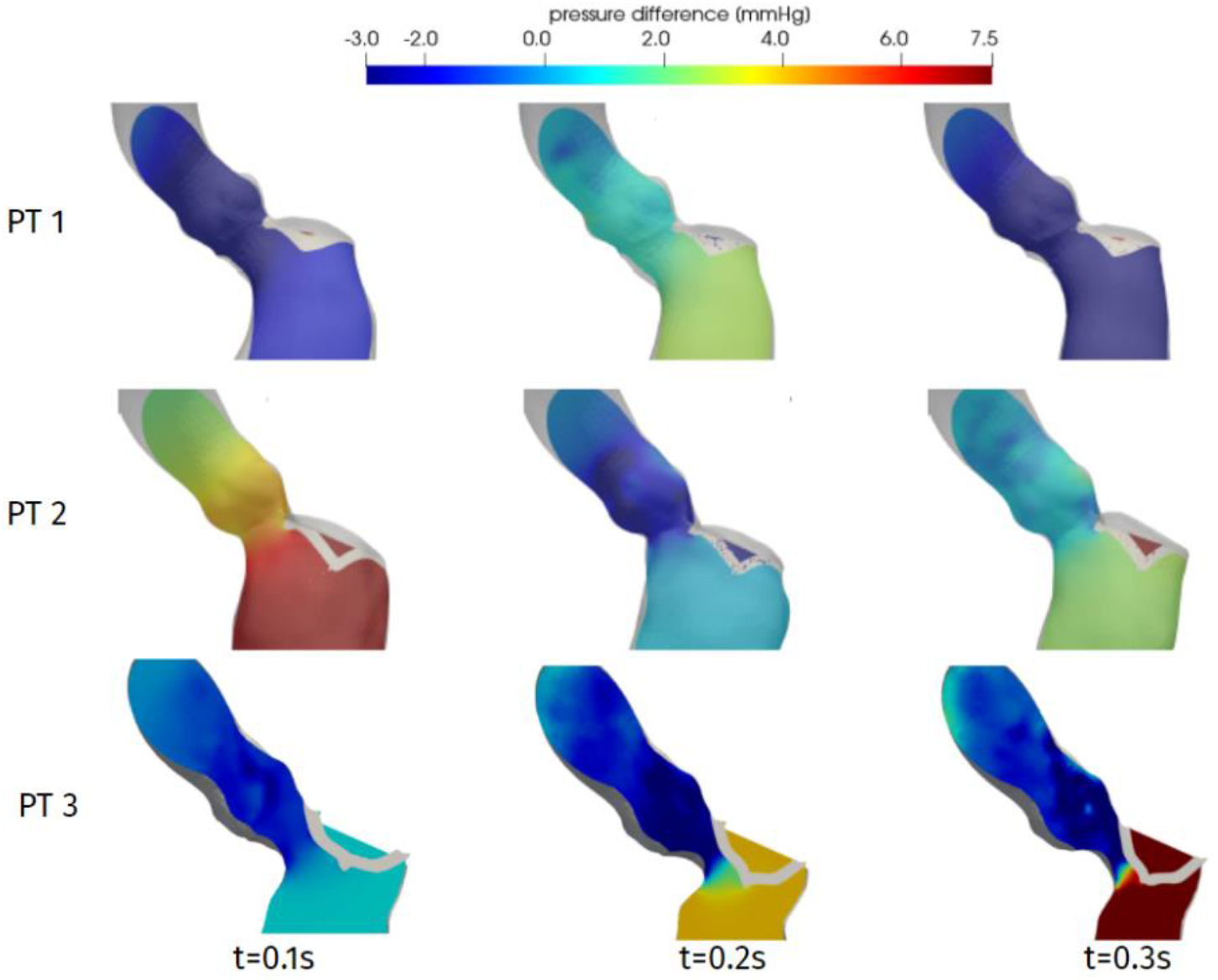
Distribution of pressure difference Δp w.r.t. sinuses, at significant times during systole.

Aiming at providing a synthetic evaluation of the severity of the hypertrophy-induced obstruction, and to provide quantitative indications on its localization and extension, in Figure 5 we present a plot of the pressure difference Δp on a line running along the septum, from the right coronary sinus of Valsalva to the LV apex. Each of these curves refers to the systolic peak of the corresponding patient, identified as the time in which the maximum velocity magnitude is attained. As a first comment, this plot confirms that the whole pressure gradient essentially develops in the LVOT and the aortic orifice. Then, we can notice that the general trend of this pressure distribution is made of a decay followed by a sudden increase that reaches the apical values of pressure in a very short length. This increase identifies the spatial position of the flow obstruction: for Patient 2 the main obstacle to the flow is represented by the aortic annulus, whereas for both Patients 1 and 3 the obstruction is within the LVOT. However, only Patient 3 has an intraventricular gradient (Δp_max_-Δp_min_=26 mmHg) that is at the limit of considering the pathological condition as hypertrophic *obstructive* cardiomyopathy, which is >30 mmHg according to the guidelines reported in [1]. Moreover, the strong pressure drop (Δp=-12 mmHg) can induce a Venturi effect that leads to the SAM of the mitral valve, as actually reported for this patient and discussed in [15].

**Figure 5.**
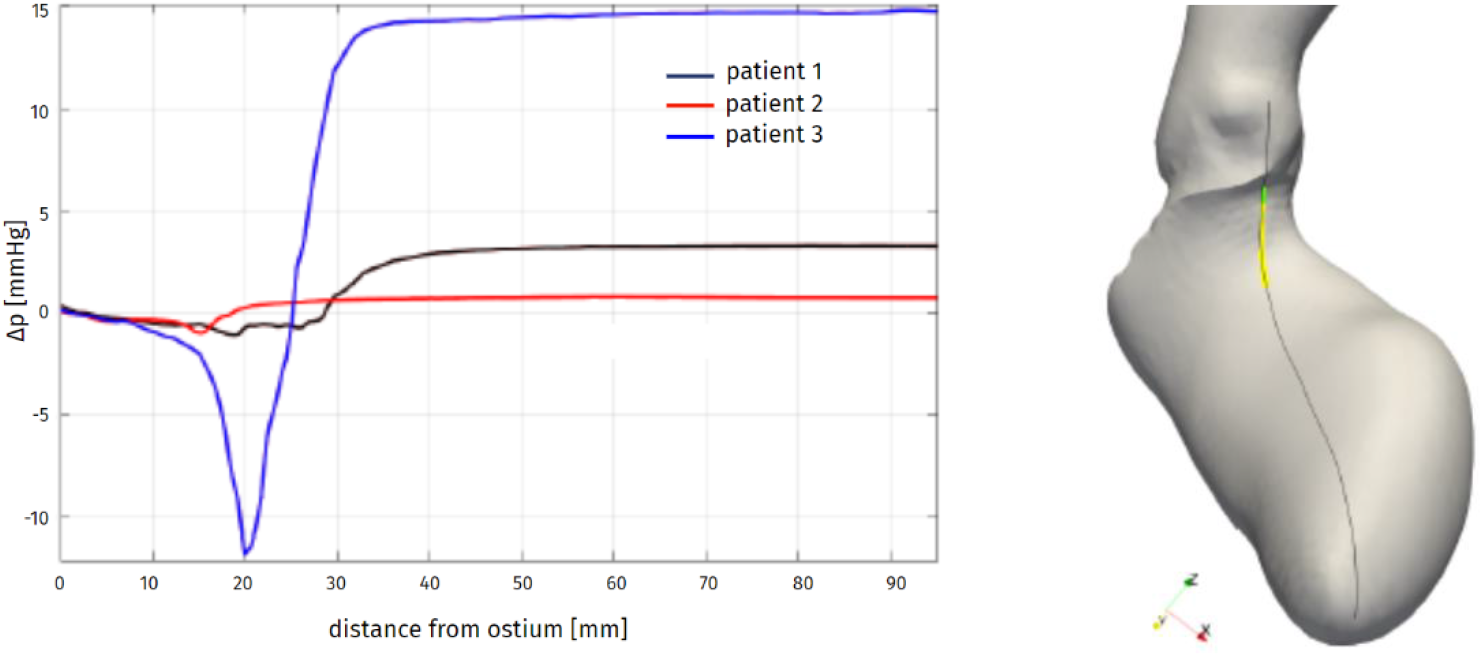
Distribution of pressure difference (left) with respect to the Valsalva sinuses along the septum (right), at the systolic peak: Patient 1, t=0.08s; Patient 2, t=0.23s; Patient 3, t=0.20s.

## DISCUSSION

We introduced and applied an image-based computational pipeline to the analysis of the hemodynamics in patients suffering from hypertrophic cardiomyopathy. Based on standard cine-MRI data regularly acquired in diagnostic procedures, with no ad-hoc acquisition series, we could create a time-dependent, volumetric artificial image with a relatively high resolution in all directions, that was then used to reconstruct the patient-specific geometry and motion of the left ventricle and ascending aorta throughout the heartbeat. The resulting time-dependent displacement was employed as a boundary condition for the computational fluid dynamics description of the blood flow, by Navier-Stokes equations in the ALE form. A resistive method was adopted to immerse the mitral valve leaflets in the domain.

The reconstruction procedure quantitatively highlights the considerable variability of the ventricle geometry and contraction among different HCM patients. In particular, different portions of the endocardium can be affected by hypokinesis, leading to different evolutions of the chamber volume and shape. The assessment the end-diastolic and end-systolic volume show how the ellipsoid approximation, commonly employed in clinical estimations, may be quite inaccurate and yield a non-negligible underestimation of the ejection fraction.

The hemodynamics results quantify the flow obstruction induced by the hypertrophy of the myocardium. The typical systolic jet through the aortic orifice has significantly higher intensity in the case of obstruction than in the other cases, and this is strongly related with the pressure gradient in the LVOT and in the Valsalva sinuses. In particular, analysing the pressure distribution along the septum, we can conclude that:

1. - The hypertrophy in Patient 1 does not induce an obstruction, since no intraventricular pressure gradient is appreciable. Nevertheless, the inhomogeneous displacement partially limits the effectiveness of the cardiac pump, with an ejection fraction that is just above the limit value of 50%.
2. - About Patient 2, we notice an intraventricular pressure gradient of small magnitude, that does not induce a pathologically intense aortic jet. However, the relatively large LV volume and rather generalized hypokinesis determine a low ejection fraction.
3. - Patient 3 definitely fits the definition of HOCM, with a strong aortic jet accompanied by a significant pressure gradient in the LVOT, and we could also quantify the Venturi effect that may have been at the origin of the development of a SAM of the mitral valve. Moreover, the obstruction has been localized, providing relevant indications for the design of a possible surgical treatment by septal myectomy.

Such conclusions demonstrate the suitability and effectiveness of the proposed computational pipeline to assess the cardiac function and the hemodynamical implications of HCM, as well as to provide clinically relevant indications for the treatment of the pathology.

## Limitations and future directions

The present study analysed only a small number of patients: investigations involving a larger set of patients may help obtaining a more comprehensive understanding of the hemodynamics of HCM, confirming the preliminary results obtained here and better accounting for the strong inter-patient variability of this condition. Moreover, a full description of the hemodynamics of this pathology should entail also a quantitative assessment of the wall shear stress oscillations – which are associated to wall cells degeneration – and of vortical structures and turbulence in the ascending aorta – that can shed further light on the effectiveness and efficiency of the cardiac pump of HCM patients. These indicators would require running several heartbeats, thus implying a significant increase in the overall computational effort. For all this reasons, a further automatization of the reconstruction procedure is currently under development, that will reduce the time that elapses between the data acquisition and the post-processing of computational results.

## Data Availability

Data is not publicly available.

## Footnotes

1 iHEART - An Integrated Heart model for the simulation of the cardiac function. European Research Council (ERC) grant agreement No 740132, P.I. Prof A. Quarteroni.

